# Based on current trends, almost all African countries are likely to report over 1 000 COVID-19 cases by the end of April 2020, and over 10 000 a few weeks after that

**DOI:** 10.1101/2020.04.05.20054403

**Authors:** Carl A. B. Pearson, Cari Van Schalkwyk, Anna M. Foss, Kathleen M. O’Reilly, SACEMA Modelling and Analysis Response Team, CMMID COVID-19 working group, Juliet R. C. Pulliam

**Affiliations:** London School of Hygiene & Tropical Medicine, London, UK; South African DSI-NRF Centre of Excellence in Epidemiological Modelling and Analysis (SACEMA), Stellenbosch University, Stellenbosch, RSA

**Keywords:** Africa, COVID-19, Computer Simulation, Epidemics

## Abstract

For African countries currently reporting COVID-19 cases, we estimate when they will report more than 1 000 and 10 000 cases. Assuming current trends, more than 80% are likely to exceed 1 000 cases by the end of April 2020, with most exceeding 10 000 a few weeks later.

## Main Text

The World Health Organization (WHO) declared COVID-19 a public health emergency of international concern (1) and then a pandemic (2), citing its rapid global spread and risk of overwhelming healthcare services with patients requiring critical care. As of 24 March 2020, WHO situation reports (SITREPs), indicated 45 African countries reported at least one laboratory-confirmed infection (“reported case”) of COVID--19 (World Health Organization 2020). Reported cases underestimate actual infections due to the mix of mild symptoms (3, 4), the similarity of symptoms common to the region (5), and weak surveillance (6). However, assuming constant reporting activity, reported cases grow in proportion to the underlying epidemic, and even with under-ascertainment of the number of actual cases, reported cases provide a useful indicator of stress on healthcare systems. We can use this surrogate for the real epidemic to forecast future trends, and understand the consequences of a slow public health response and what preparations need to be made now.

## The Study

We use a branching process model to project the number of future cases of COVID-19 in each country. This model assumes each case produces a number of new cases (distributed *N*∼NegBinom(*R* = 2, *k* = 0.58)) (7, 8) which occur after some delay (distributed *T*∼LogNormal(E[*X*] = 4.7, SD[*X*] = 2.9) (9). We start with cases corresponding to the first 25 (or fewer) reported cases in the WHO SITREPs up to 23 March 2020 (10). Using those epidemic parameters and initial cases and dates, we simulate the accumulation of the reported cases. We assume there are always sufficient unreported infections to continue transmission, and that new cases represent a reporting sample from both identified and unidentified transmission chains. As long as a constant reporting fraction persists during this period, and unreported spread is large relative to reported cases (or reporting has negligible impact on control), this is a reasonable approximation.

For each set of country-specific initial conditions, we generate n=10 000 epidemics, discarding any that fade out, consistent with our assumption of unreported transmission chains. We identify the dates when each simulation run crosses 1 000 and 10 000 reported cases, and then evaluate the 50% and 95% quantiles of those dates to determine the forecast interval. The model was built in the R statistical programming language, using the *bpmodels* package (11), and the *data2019nCoV* package for the SITREP data (12). All analysis code is available from https://github.com/SACEMA/COVID10k.

We project that almost all African countries are likely to pass 1 000 reported cases by the end of April 2020 and 10 000 within another few weeks (Figure 1 and Table 1); alarmingly, these are largely synchronized continent-wide and real disease burden will certainly exceed reported cases. Since our projections assume failed containment of initial cases and no interventions reducing early transmission, they are pessimistic relative to any benefits of local action. However, containment measures, e.g. travel restrictions, increased testing, contact tracing, isolation of cases and quarantine of contacts, are likely to slow, but not halt, real epidemic growth (13). Indeed, increased testing may accelerate the time to reporting these numbers, as improved ascertainment increases the identified fraction of real cases. However, the model also optimistically assumes surveillance capacity is not overwhelmed or stymied, which would slow reaching 1 000 reported cases while the real disease burden grows uncontrolled. Because we ignore these effects, the model is only appropriate for short-range forecasts.

**Table 1.**
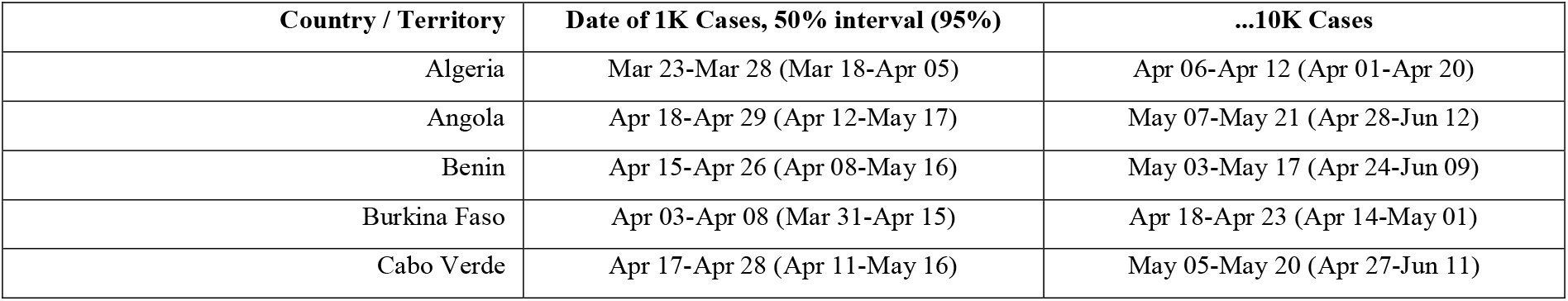
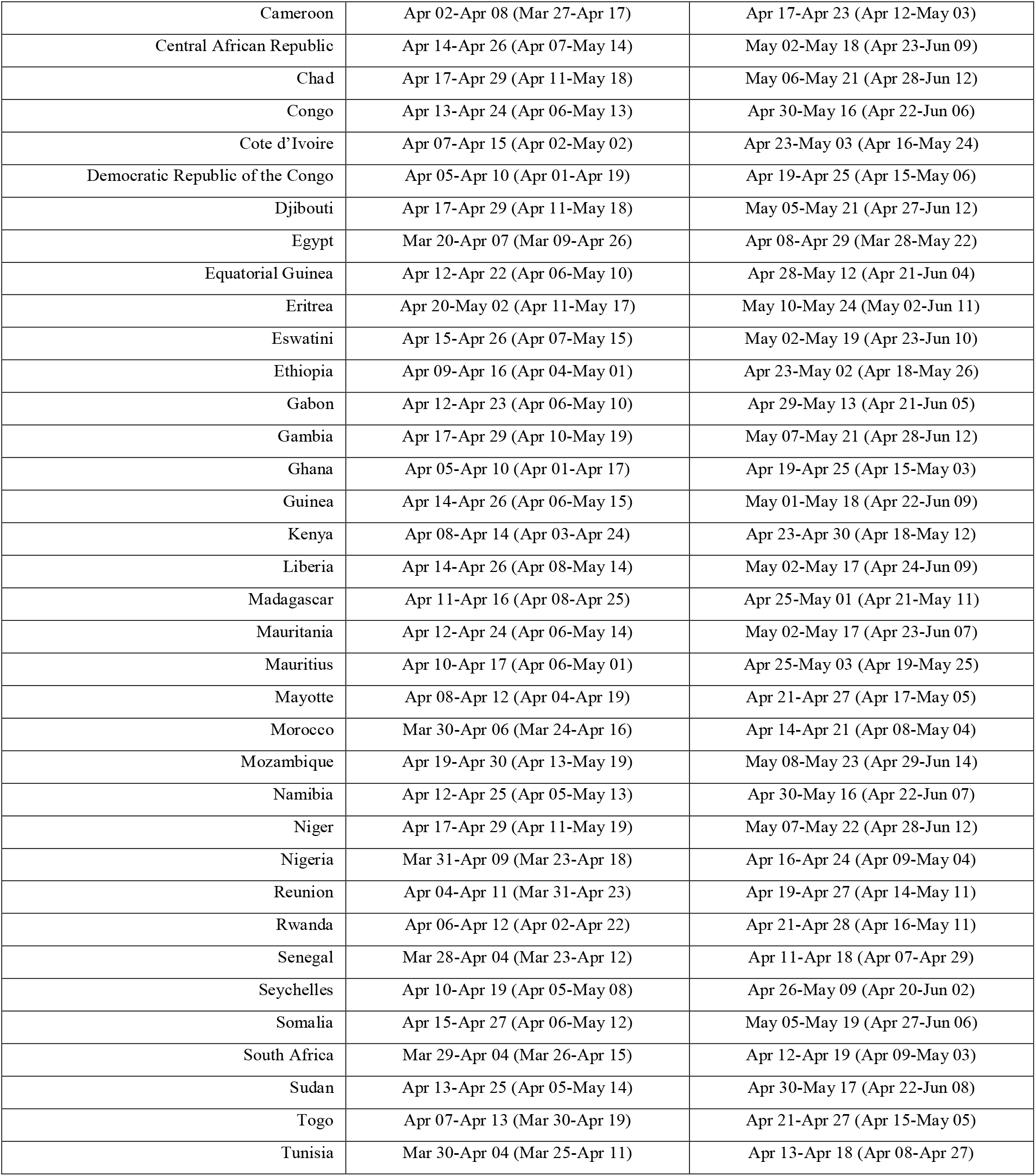
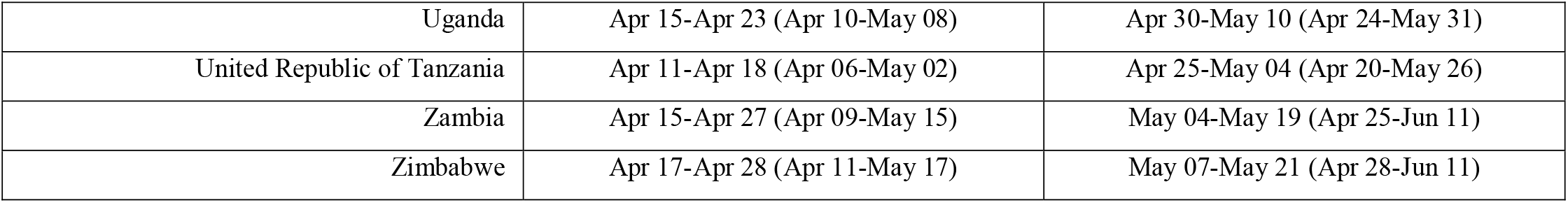
Projected Timing of Reporting 1 000 and 10 000 COVID-19 cases for all African Countries Reporting Cases as of 23 March 2020.

| Country / Territory | Date of 1K Cases, 50% interval (95%) | ...10K Cases |
| --- | --- | --- |
| Algeria | Mar 23-Mar 28 (Mar 18-Apr 05) | Apr 06-Apr 12 (Apr 01-Apr 20) |
| Angola | Apr 18-Apr 29 (Apr 12-May 17) | May 07-May 21 (Apr 28-Jun 12) |
| Benin | Apr 15-Apr 26 (Apr 08-May 16) | May 03-May 17 (Apr 24-Jun 09) |
| Burkina Faso | Apr 03-Apr 08 (Mar 31-Apr 15) | Apr 18-Apr 23 (Apr 14-May 01) |
| Cabo Verde | Apr 17-Apr 28 (Apr 11-May 16) | May 05-May 20 (Apr 27-Jun 11) |
| Cameroon | Apr 02-Apr 08 (Mar 27-Apr 17) | Apr 17-Apr 23 (Apr 12-May 03) |
| Central African Republic | Apr 14-Apr 26 (Apr 07-May 14) | May 02-May 18 (Apr 23-Jun 09) |
| Chad | Apr 17-Apr 29 (Apr 11-May 18) | May 06-May 21 (Apr 28-Jun 12) |
| Congo | Apr 13-Apr 24 (Apr 06-May 13) | Apr 30-May 16 (Apr 22-Jun 06) |
| Cote d'Ivoire | Apr 07-Apr 15 (Apr 02-May 02) | Apr 23-May 03 (Apr 16-May 24) |
| Democratic Republic of the Congo | Apr 05-Apr 10 (Apr 01-Apr 19) | Apr 19-Apr 25 (Apr 15-May 06) |
| Djibouti | Apr 17-Apr 29 (Apr 11-May 18) | May 05-May 21 (Apr 27-Jun 12) |
| Egypt | Mar 20-Apr 07 (Mar 09-Apr 26) | Apr 08-Apr 29 (Mar 28-May 22) |
| Equatorial Guinea | Apr 12-Apr 22 (Apr 06-May 10) | Apr 28-May 12 (Apr 21-Jun 04) |
| Eritrea | Apr 20-May 02 (Apr 11-May 17) | May 10-May 24 (May 02-Jun 11) |
| Eswatini | Apr 15-Apr 26 (Apr 07-May 15) | May 02-May 19 (Apr 23-Jun 10) |
| Ethiopia | Apr 09-Apr 16 (Apr 04-May 01) | Apr 23-May 02 (Apr 18-May 26) |
| Gabon | Apr 12-Apr 23 (Apr 06-May 10) | Apr 29-May 13 (Apr 21-Jun 05) |
| Gambia | Apr 17-Apr 29 (Apr 10-May 19) | May 07-May 21 (Apr 28-Jun 12) |
| Ghana | Apr 05-Apr 10 (Apr 01-Apr 17) | Apr 19-Apr 25 (Apr 15-May 03) |
| Guinea | Apr 14-Apr 26 (Apr 06-May 15) | May 01-May 18 (Apr 22-Jun 09) |
| Kenya | Apr 08-Apr 14 (Apr 03-Apr 24) | Apr 23-Apr 30 (Apr 18-May 12) |
| Liberia | Apr 14-Apr 26 (Apr 08-May 14) | May 02-May 17 (Apr 24-Jun 09) |
| Madagascar | Apr 11-Apr 16 (Apr 08-Apr 25) | Apr 25-May 01 (Apr 21-May 11) |
| Mauritania | Apr 12-Apr 24 (Apr 06-May 14) | May 02-May 17 (Apr 23-Jun 07) |
| Mauritius | Apr 10-Apr 17 (Apr 06-May 01) | Apr 25-May 03 (Apr 19-May 25) |
| Mayotte | Apr 08-Apr 12 (Apr 04-Apr 19) | Apr 21-Apr 27 (Apr 17-May 05) |
| Morocco | Mar 30-Apr 06 (Mar 24-Apr 16) | Apr 14-Apr 21 (Apr 08-May 04) |
| Mozambique | Apr 19-Apr 30 (Apr 13-May 19) | May 08-May 23 (Apr 29-Jun 14) |
| Namibia | Apr 12-Apr 25 (Apr 05-May 13) | Apr 30-May 16 (Apr 22-Jun 07) |
| Niger | Apr 17-Apr 29 (Apr 11-May 19) | May 07-May 22 (Apr 28-Jun 12) |
| Nigeria | Mar 31-Apr 09 (Mar 23-Apr 18) | Apr 16-Apr 24 (Apr 09-May 04) |
| Reunion | Apr 04-Apr 11 (Mar 31-Apr 23) | Apr 19-Apr 27 (Apr 14-May 11) |
| Rwanda | Apr 06-Apr 12 (Apr 02-Apr 22) | Apr 21-Apr 28 (Apr 16-May 11) |
| Senegal | Mar 28-Apr 04 (Mar 23-Apr 12) | Apr 11-Apr 18 (Apr 07-Apr 29) |
| Seychelles | Apr 10-Apr 19 (Apr 05-May 08) | Apr 26-May 09 (Apr 20-Jun 02) |
| Somalia | Apr 15-Apr 27 (Apr 06-May 12) | May 05-May 19 (Apr 27-Jun 06) |
| South Africa | Mar 29-Apr 04 (Mar 26-Apr 15) | Apr 12-Apr 19 (Apr 09-May 03) |
| Sudan | Apr 13-Apr 25 (Apr 05-May 14) | Apr 30-May 17 (Apr 22-Jun 08) |
| Togo | Apr 07-Apr 13 (Mar 30-Apr 19) | Apr 21-Apr 27 (Apr 15-May 05) |
| Tunisia | Mar 30-Apr 04 (Mar 25-Apr 11) | Apr 13-Apr 18 (Apr 08-Apr 27) |
| Uganda | Apr 15-Apr 23 (Apr 10-May 08) | Apr 30-May 10 (Apr 24-May 31) |
| United Republic of Tanzania | Apr 11-Apr 18 (Apr 06-May 02) | Apr 25-May 04 (Apr 20-May 26) |
| Zambia | Apr 15-Apr 27 (Apr 09-May 15) | May 04-May 19 (Apr 25-Jun 11) |
| Zimbabwe | Apr 17-Apr 28 (Apr 11-May 17) | May 07-May 21 (Apr 28-Jun 11) |

**Figure 1.**
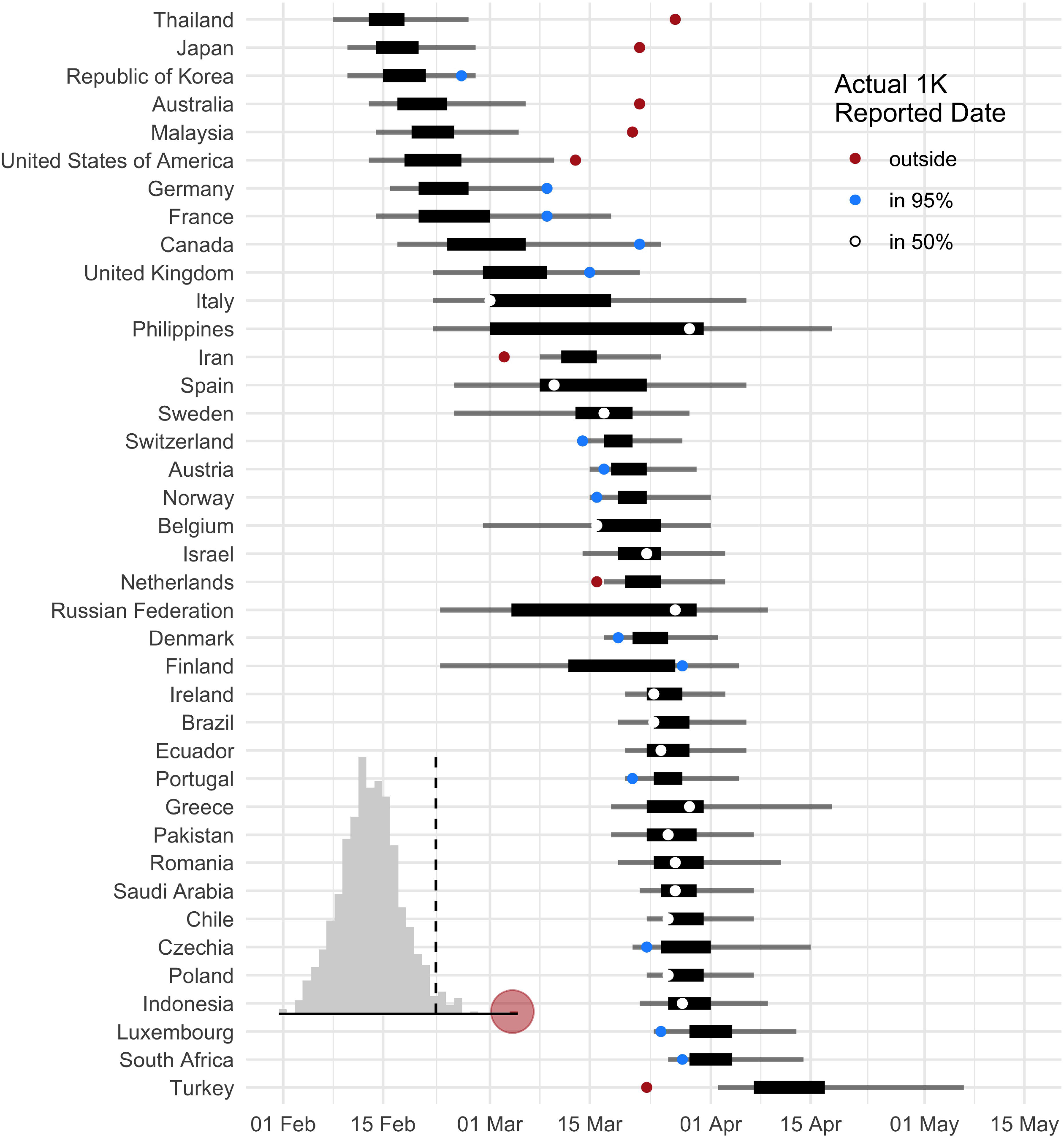
Distribution of times to 1 000 (red to yellow) and 10 000 (grey) cases, with inset map indicating median expected arrival dates by country (red to yellow, corresponding to time distributions; countries not reporting cases by 23 March 2020 SITREP in grey).

As model validation, we applied this same forecasting approach to countries world-wide that have now exceeded 1 000 reported cases; we did not consider those with more than 10 000 cases, as they have all undergone substantial control measures modifying epidemic growth. We found that 44% of actual reporting dates fell within the 50% prediction intervals, and 79% within the 95% interval (Figure 2), indicating the forecast prediction interval is too certain, as expected for a rapid but low detail model. We further showed that forecast performance is not a random outcome by performing a randomization test: we shuffle the assignment of forecast days-to-1 000-cases to different countries, and score 1 000 shuffled predictions; the real forecast score is significantly different from random at the p < 0.001 level (Figure 2 inset).

**Figure 2.**
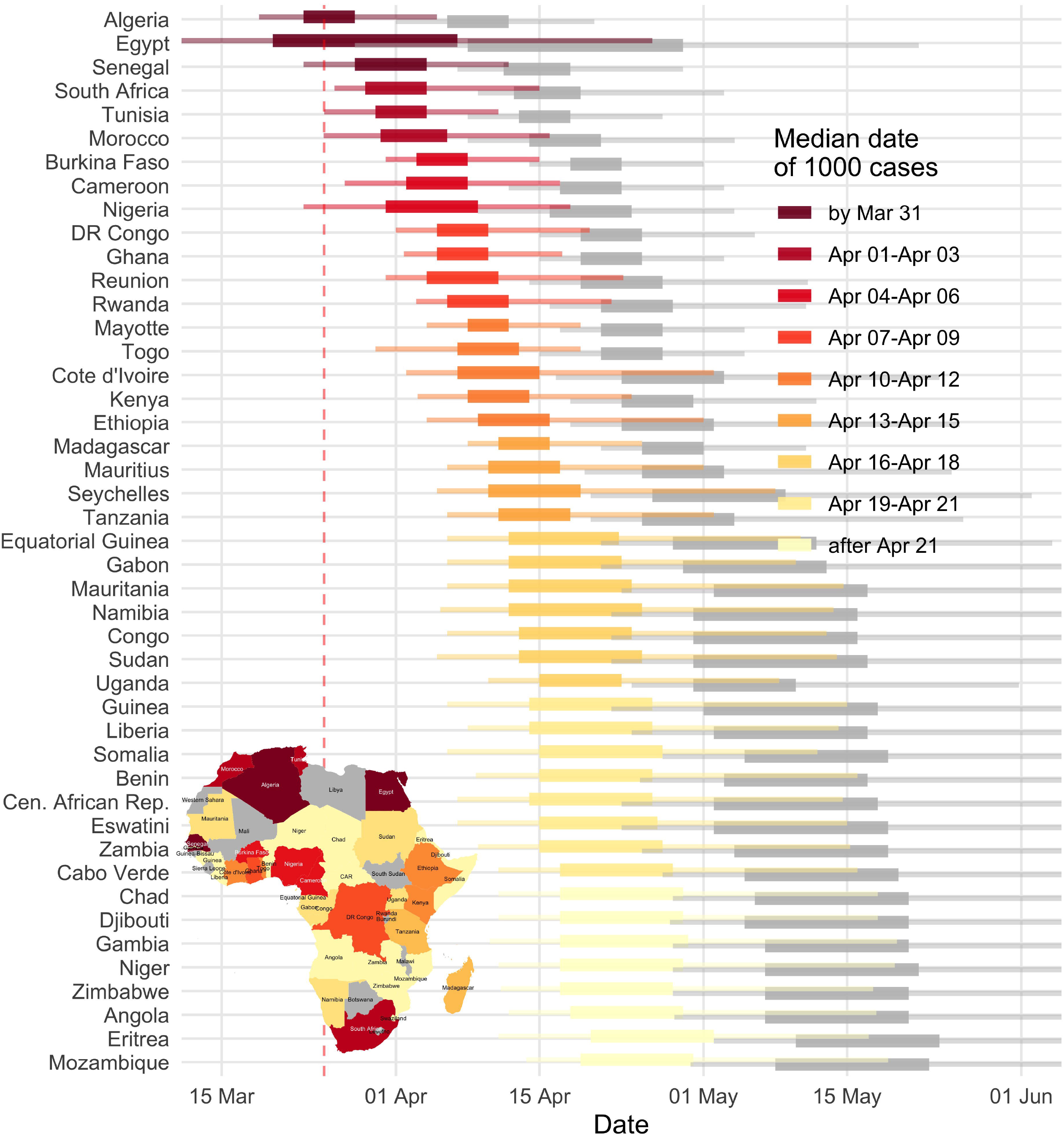
Forecast validation for countries having already reached 1 000 cases; inset distribution indicates randomization results for actual forecast (red) versus randomly assigned forecast (grey), with 0.975 quantile indicated by the line. For actual reporting dates, 44% fell within the 50% prediction intervals, and 79% within the 95%.

Specific to Africa, the forecast for South Africa fell within the 50% prediction interval (SITREP 69; 29 March 2020). From 23 March 2020, we projected a few other countries would also likely be crossing this threshold soon: Algeria, Egypt, Morocco, Senegal and Tunisia. As of SITREP 75 (4 April 2020), the first three are still fast approaching this limit, while fast and intense responses in the latter two may have successfully slowed the epidemic.

## Conclusions

Using reporting to date, and assuming similar epidemiological trends to those seen globally, we project that almost all African countries are likely to exceed 1 000 reported cases by the end of April 2020, and 10 000 within another few weeks. This timing is largely synchronized continent-wide and real disease burden will certainly exceed reported cases. Our projections assume no substantive changes between the initially reported cases and the forecast points; while some countries have taken drastic actions, many have not or have acted slowly. As seen in other regions, because onset of severe symptoms can be delayed weeks from infection and last several weeks, interventions have limited immediate impact on new hospitalizations or facility demand, meaning that most of the countries in our projection would be well past 1 000 cases by the time the effects of interventions started today would be observed.

These results call for accelerated preparations across Africa to ready healthcare systems and citizens for the incoming wave of COVID-19 infections. Augmented staffing, personal protective equipment stores and training, general isolation beds and equipped critical care units are all urgently needed. Citizen awareness will also be critical, and officials should encourage preventive measures such as physical distancing and regular hand washing.

## Data Availability

All model code and data are available from https://github.com/SACEMA/COVID10k

https://github.com/eebrown/data2019nCov

## Acknowledgments

CABP gratefully acknowledges funding of the NTD Modelling Consortium by the Bill and Melinda Gates Foundation (OPP1184344). KMO gratefully acknowledges funding of the Effectiveness of Supplementary Immunization Activities by the Bill and Melinda Gates Foundation (OPP1191821).

The Centre for Mathematical Modelling of Infectious Disease 2019-nCoV working group includes: Emily S Nightingale, Sebastian Funk, Rosalind M Eggo, Joel Hellewell, Adam J Kucharski, Quentin J Leclerc, Nicholas G. Davies, Jon C Emery, Stefan Flasche, Nikos I Bosse, Sam Abbott, Megan Auzenbergs, Amy Gimma, Simon R Procter, Rein M G J Houben, Timothy W Russell, Akira Endo, Charlie Diamond, James D Munday, Gwen Knight, Fiona Yueqian Sun, Yang Liu, Arminder K Deol, Thibaut Jombart, Billy J Quilty, Samuel Clifford, Petra Klepac, Kevin van Zandvoort, Kiesha Prem, Alicia Rosello, Graham Medley, Mark Jit, Christopher I Jarvis, Hamish Gibbs, and W John Edmunds.

The SACEMA Modelling and Analysis Response Team (SMART) includes: Roxanne Beauclair, Elisha B. Are, Olatunji O. Adetokunboh, Jeremy Bingham, C. Marijn Hazelbag, Ivy Kombe, and Joseph B. Sempa.

## Author Bio

Carl A. B. Pearson is a Research Fellow at the London School of Hygiene & Tropical Medicine, and a Research Fellow with the South African DSI-NRF Centre of Excellence in Epidemiological Modelling and Analysis (SACEMA) at Stellenbosch University. His primary research focus is modelling infectious disease dynamics to understand the optimal evaluation and application of interventions, particularly vaccines.

